# Factors associated with adherence to HIV testing guidelines among HIV negative female sex workers in Kampala Uganda

**DOI:** 10.1101/2022.01.18.22269503

**Authors:** Lydia Atuhaire, Constance S Shumba, Lovemore Mapahla, Innocent Maposa, Peter S Nyasulu

## Abstract

**Background:** Frequent HIV testing at intervals of every three to six months is recommended among HIV negative female sex workers as the first entry point to HIV prevention and treatment. In this study, we examined the extent to which HIV negative female sex workers adhere to the testing guidelines by measuring the frequency of testing in the last 12 months and identified associated factors among female sex workers in Kampala Uganda.

**Methods:** We conducted a cross-sectional study using structured questionnaires. Using equal probability selection method, 12 hotspots were selected, and 200 participants interviewed based on proportional allocation in each hotspot. We used descriptive statistics to describe female sex workers’ characteristics, and multivariable logistic regression model to determine the factors associated with their adherence to the HIV testing guidelines. Adherence to the HIV testing guidelines was defined as having tested three or more times in the last 12 months. Factors with p-value ≤0.05 significance level were considered statistically significant.

**Results:** From the 200 study participants, 43% were aged 25-30 years, joined sex work between the ages of 18 to 24 years, 49% had attained primary and 41% secondary school respectively. 88% reported HIV testing status, of which 56% had tested three or more times in the 12 months preceding the survey. Attaining secondary education, was independently associated with adherence to the HIV testing guidelines (OR 1.86, 95% CI: 1.01 - 3.44, p=0.047). Those that had tested for STIs in the last three months (OR= 2.13, 95% CI: 0.95 - 4.74, p=0.065) and accessed HIV testing in a drop-in centre (OR= 5.90, 95% CI: 0.71 - 49.1, p=0.101) had higher odds of adhering to the HIV testing guidelines

**Conclusion:** This study found suboptimal adherence to the HIV testing guidelines among HIV negative female sex workers. Interventions such as HIV self-testing that improve access to, and frequency of HIV testing need to be taken to scale. There is need for rigorous behaviour change program evaluation to continually refine understanding of the message mix, to ensure simplified and easy to comprehend awareness messages are designed for female sex workers with no formal education.

## Introduction

Globally, female sex workers (FSWs) are recognised as a population that is at increased risk of acquiring HIV infection. As such this population is a critical target for frequent HIV testing, an essential component of combination HIV prevention interventions (1). Early HIV testing is the first entry to HIV prevention and treatment which places an emphasis on the high risk groups in the population such as FSWs (2). The World Health Organization (WHO) recommends frequent HIV testing at intervals of every three months to six months among FSWs, and countries are at liberty to adapt a period within that timeframe that suits their population and epidemic (3). The WHO rolled out guidelines and implementation approaches to be used to support access to client centred routine HIV testing, care and prevention services to reduce continued transmission of HIV infection (3, 4). Countries in sub-Saharan Africa adapted the WHO HIV testing policies released in 2015 and in Uganda, specific testing guidelines released in 2016 indicate that FSWs should test every three months (5, 6)

Various approaches have been operationalised and are being implemented to improve routine HIV testing among FSWs. Such interventions include: integration of HIV testing services (7) with other health services such as family planning (8), community based testing within the sex work hotspots in Uganda and Kenya; and HIV testing in drop-in-centres in the Democratic Republic of Congo (9-11), community mapping for testing in South Africa (12) as well as several other community-based testing interventions within the broader sub Saharan Africa region (13-16). Successes of these interventions have been reported such as ‘sex work hotspot-based testing’ which has been shown to have potential for reducing undiagnosed HIV from 71.6% to 20.2% among FSWs and young women (11). Despite these successes, the desired uptake of routine HIV testing among FSWs is still suboptimal (17, 18).

A study conducted in Benin (19) reported a 40% HIV testing uptake among FSW in the last three months, and only 21% had tested in the last 3 to 6 months. A cross-sectional survey that measured HIV testing at different time points among FSWs in Mombasa, Kenya found that 45% were testing for HIV infection every 3 months, 15% every 6 months while only 12% had tested in the last 1 to 12 months (20). A few studies that found high levels of testing among FSWs, that is, 89.2% (21) and 88% (20) used a different measuring approach. The investigators measured longer time periods between testing, that is, at 12 months, and ever tested in a lifetime.

Consistent with other countries in sub-Saharan Africa, the uptake of routine HIV testing among FSWs is low in Uganda. A previous study conducted among FSWs in Kampala showed that only 53% of FSWs had ever tested for HIV, and of these, 16% had tested in the preceding 12 months (22). A cross-sectional survey conducted in 2019 which assessed FSWs preference and uptake of different community-based HIV testing service delivery models found that 86% FSWs had taken an HIV test in the last 12 months (9). A related study that assessed HIV screening practices among FSWs in the four regions of Uganda showed 95% had ever tested for HIV and 67% had tested for HIV two or more times in the last 12 months preceding the survey (23).

HIV testing among FSWs is propagated by several factors which have previously been reported to influence routine HIV testing. These include behavioural factors such as high alcohol intake, drug use, and high mobility of FSWs in search of new customers (18, 20, 21). Further, there are structural factors such as stigma and discrimination, violence and violation of rights (24, 25), and health system factors such as the limited availability of trained health workers to provide friendly services, inadequate friendly testing centres due to conflicting hours of testing with FSWs schedules of work. This acts as a catalyst for FSWs to be reluctant to seek routine HIV testing services (26-28). Programs that have attempted to understand the contextual issues that hinder access to HIV services among FSWs have led to scale up of comprehensive and targeted FSWs approaches. These program strategies have also registered great success in improving access to the HIV services which include routine HIV testing and early diagnosis of HIV infection (29). For example, the enhanced peer outreach approach that targeted key populations including FSWs in Cote d’Ivoire, the Democratic Republic of Congo, and Burundi, successfully identified and offered HIV testing to the hard-to-reach FSWs. This approach led to an increase in diagnoses of new HIV infection among FSWs, and improved linkage to care and treatment (29, 30).

Although studies in sub Saharan Africa and in particular Uganda (22, 23) have studied uptake of HIV testing, the majority have focused at examining the history of ever tested, and how proximate the tests were (18, 21, 24, 26, 31). These studies did not assess the extent FSWs adhered to the HIV testing guidelines (3). Limited data are available regarding the frequency and factors associated with routine HIV testing among FSWs in Uganda. This study therefore examines the extent to which HIV negative FSWs adhere to the testing guidelines by measuring the frequency of testing in the previous year. It also assesses the factors associated with adherence to HIV testing guidelines among HIV negative FSWs.

## Methods

### Ethics consideration

The study was approved by the Institutional Review Boards of the Uganda Virus Research Institute (UVRI) reference number GC/12719/08/723, and Ethics Committee of Faculty of Medicine and Health Sciences of Stellenbosch University reference number S19/05/088, and Uganda National Council of Science and Technology reference number HS-2665. All participants were given detailed information about the study and were informed that their participation was voluntary and that they had the right to withdraw from the interviews at any time. Participants also provided verbal informed consent as opposed to written consent, in line with the guidance from the Uganda National Council of Science and Technology (reference number HS-2665) on conducting research among key populations.

### Study design

We conducted a cross-sectional study using structured questionnaires. We used simple random sampling to select the hotspots and non-probability sampling methods specifically voluntary response sampling to select participants, we assessed adherence to the HIV testing guidelines for FSWs by determining the number of times FSWs tested for HIV in the last 12 months preceding the study

### Study setting

We conducted the study at established sex work hotspot areas in the five administrative divisions of Kampala city. We defined these sex work hotspots as geographical locations where the actual sex work trade is concentrated such as streets, lodges, and bars. Kampala is the capital and largest city in Uganda. The city is divided into five administrative divisions from from where the sample was drawn, namely, Nakawa, Makindye, Kampala Central, Rubaga and Kawempe divisions. We selected Kampala because it is a highly active and economically vibrant business centre and attracts potential customers for FSWs due to financial accessibility. The city has many truck drivers, that transport commercial goods across Kampala from neighbouring countries of Rwanda, Democratic Republic of the Congo, Tanzania, Kenya, Burundi, Rwanda, and South Sudan.

### Study population

The study population consisted of FSWs aged ≥18 years who operated in any of the sex work hotspots within any of the five administrative divisions of Kampala at the time of the study. We recruited FSWs who were HIV negative only based on self-report. We defined FSWs as individuals who offer sex in exchange for money or other financial or material benefits as their main source of income generation or supplements other primary source of income, with sex work as an additional source of income. We recruited individuals engaged in this trade and had been operating within the last 12 months preceding the survey.

### Variable measurements

The main outcome measure was adherence to the HIV testing guidelines for FSWs. Adherence was defined as having tested for HIV three times or more within the last 12 months. The exposure variables were demographic and social behavioural characteristics including age, education level, marital status, housing status, and if the FSW had other job other than sex work. The social behavioural exposure variables are indicated in table 2.

**Table 1:** Socio-demographic characteristics of the study participants.

| **Socio-demographic characteristics** | **Frequency n=200** | **Percentage** |
| --- | --- | --- |
| **Location** |  |  |
| Kampala Central | 60 | 30.0 |
| Nakawa | 22 | 11.0 |
| Kawempe | 62 | 31.0 |
| Rubaga | 37 | 18.5 |
| Makindye | 19 | 9.5 |
| **Age group** |  |  |
| 18-24 | 58 | 29.0 |
| 25-30 | 85 | 42.5 |
| 31-49 | 57 | 28.5 |
| **Age at sex work start** |  |  |
| 10-17 | 28 | 14.2 |
| 18-24 | 100 | 50.8 |
| 25-45 | 69 | 35.0 |
| Missing | 3 | 1 |
| **Education** |  |  |
| None | 18 | 9.0 |
| Primary | 97 | 48.5 |
| Secondary | 82 | 41.0 |
| Post-Secondary | 3 | 1.5 |
| **Marital Status** |  |  |
| Single/Never married | 44 | 22.0 |
| Married and cohabiting | 18 | 9.0 |
| Separated/Divorced/Widowed | 138 | 69.0 |
| **Has children** |  |  |
| No | 28 | 14 |
| Yes | 171 | 85.5 |
| Missing | 1 | 0.5 |
| **Other Job other than sex work** |  |  |
| No | 128 | 64 |
| Yes | 71 | 35.5 |
| Missing | 1 | 0.5 |
| **Housing status n=124** |  |  |
| Own/rented house | 178 | 89.0 |
| Housed by relatives/friends | 18 | 9.0 |
| No shelter | 4 | 2.0 |

**Table 2:** Characteristics of FSWS by adherence to the HIV testing guidelines status.

| Variable | Non adherence<br>N (%) | Adherence<br>N (%) | p-value |
| --- | --- | --- | --- |
| **Age group (Age n=179)** |  |  |  |
| 18-24 | 22(27.9) | 33(33.0) | 0.398 |
| 25-30 | 38(48.1) | 38(38.0) |  |
| 31-49 | 19(24.1) | 29(29.0) |  |
| **Education n=179** |  |  |  |
| None | 30(37.9) | 32(32) | 0.03 |
| Primary | 20(25.3) | 15(15) |  |
| ≥Secondary | 29(36.7) | 53(53) |  |
| **Marital Status\* n=179** |  |  |  |
| Single/Never married | 15(19.0) | 26(26.0) | 0.512 |
| Married/cohabiting | 9(11.4) | 9(9.0) |  |
| Separated/Divorced/Widowed | 55(69.6) | 65(65.0) |  |
| **Other Jobs n=178** |  |  |  |
| No | 50(63.3) | 63(63.6) | 0.962 |
| Yes | 29(36.7) | 36(36.4) |  |
| **Housing status n=124** |  |  |  |
| Own/rented house | 67(84.8) | 91(91.0) | 0.296 |
| Housed by relatives/friends | 11(13.9) | 7(7.0) |  |
| No shelter | 1(1.3) | 2(2.0) |  |
| **Condom use by FSW n=179** |  |  |  |
| Consistence | 10(12.7) | 11(11.0) | 0.732 |
| Inconsistence | 69(87.3) | 89(89.0) |  |
| **Drug use (Ever used) n=176** |  |  |  |
| No | 58(74.4) | 64(65.3) | 0.196 |
| Yes | 20(25.6) | 34(34.7) |  |
| **Regular medical check-up n=179** |  |  |  |
| No | 59(74.7) | 64(64) | 0.126 |
| Yes | 20(25.3) | 36(33) |  |
| **Knowledge on testing guidelines n=178** |  |  |  |
| No | 24(30.8) | 40(40) | 0.203 |
| Yes | 54(69.2) | 60(33) |  |
| **Most recent HIV testing n=179** |  |  |  |
| No | 57(72.1) | 73(73) | 0.899 |
| Yes | 22(27.8) | 27(27) |  |
| **Social support for HIV test n=178** |  |  |  |
| Yes | 36(45.6) | 48(48.5) | 0.699 |
| No | 43(54.4) | 51(51.5) |  |
| **Receipt of minimum health package in last 3 months n=179** |  |  |  |
| No | 79(100) | 82(82.0) | P<0.001 |
| Yes | 0(33) | 18(18.0) |  |
| **HIV testing awareness campaign n=179** |  |  |  |
| No | 12(15.2) | 10(10.0) | 0.294 |
| Yes | 67(84.8) | 90(90.0) |  |
| **Concerns about stigma and discrimination n=178** |  |  |  |
| High | 31(39.7) | 36(36.0) | 0.567 |
| Moderate | 20(25.6) | 33(33.0) |  |
| Low | 27(34.6) | 31(31.0) |  |
| **FSW risk perception to HIV risk n=176** |  |  |  |
| No risk | 11(14.5) | 14(14.0) | 0.747 |
| low risk | 33(43.4) | 49(49.0) |  |
| ≥High risk | 32(42.1) | 37(37.0) |  |

### Sample size and sampling procedure

We first obtained a list of sex work hotspots with an estimated population of FSWs from the Infectious Diseases Institute. This institute works with FSW network organizations in Kampala hence has more accurate data of FSWs population in the city. There were 327 mapped hotspots with an estimated total number of 11,558 FSWs operating in those hotspots. Using equal probability selection method of the hotspots, 12 sites were randomly selected from the five divisions, and these yielded a total of 6,761 FSWs. Due to resource constraints, out of the 6,761 FSWs in 12 hotspots, a final sample of 200 was selected basing on proportional allocation in each hotspot.

### Participant recruitment

We approached the FSWs network organisations, introduced the study, and asked them to help us identify the FSW peer leaders from the selected sex work hotspots. We conducted an introductory session with the FSW peer leaders and oriented them on to the objectives of the study, and all issues related to ethical conduct of the survey. We emphasized the confidentiality, safety, privacy, and voluntary nature of their participation in the study. After orientation to the study, the peer leaders approached various FSWs and introduced them to the study. Those who expressed willingness to participate were allowed to identify a mutually agreed private and safe place for a detailed interview. This recruitment process was conducted on a first come, first served basis until the number for each hotspot was reached.

### Data collection procedures

#### Validation of tools and data quality assurance

We developed a structured questionnaire and consenting documents based on the empirical literature, and prior experience of the research team in HIV programming. From the comprehensive review of literature, potential factors that influence adherence to the HIV testing guidelines were categorized under different domains. The domains were social behavioral practices, health seeking behavior, HIV testing, availability and access to HIV prevention services, risk perception and stigma and discrimination and depression tendencies. After the key domains were identified, relevant questions relating to each domain were developed by considering Uganda and FSWs contexts. Accordingly, 98 questions assessing all the domains on adherence to testing guidelines were initially developed. Two members of the team with expert knowledge in HIV prevention did content validation of the tool to select the most relevant and reduced the total number to 60. The team also assessed clarity of the instrument. Discussion on each question with two people of non-medical background was also made to further ensure clarity and face validity of the instrument.

The tools were translated to Luganda, a common local language in Kampala and back translated into English. The questionnaire was piloted in a non-study sex work hotspot in Wakiso district neighboring Kampala city. A total of 12 FSWs were enrolled in the pilot testing process. The pilot testing of the questionnaire helped to determine the limitations of the survey regarding the format of the questions clarity, appropriate wording, and the estimate time of completion per each questionnaire. The pilot testing was stopped after the observed limitations in the questionnaire were corrected.

### Data collection

Data were collected using a hard copy structured questionnaire administered by interviewers in one-on-one interviews. A research team with experience in quantitative data collection was recruited and trained to enhance their knowledge in quantitative data collection practices and research ethics, so that they could adhere to the protection of participant’s privacy, confidentiality, safety, and all other ethics standards. FSWs who provided verbal informed consent were interviewed in a private place mutually agreed on with the researcher. Data collection was done between November to December 2020.

### Data management

Data quality checks were conducted daily during field monitoring by crosscheck discrepancies, and completeness of data on all variables. We conducted real time form review and corrected missing or wrongly captured data while still in the field when it was still possible to access participants. Data were entered in Research Electronic Data Capture (REDCap), an online data management tool for surveys which helps to quickly find discrepancies and errors in the collected data (32). Data were exported into Stata version 15.0 where further checking for errors, outliers, and completeness were conducted.

### Statistical analysis

The frequencies and corresponding percentages were used to describe categorical variables. Only complete cases were used in the analysis of the dataset. Univariate logistic regression models were done for each independent variable and the binary study outcome, that is, adherence to HIV testing guidelines among HIV-negative female sex workers. All variables from univariate models, which had a p-value of 0.2 were included in subsequent multivariable model. Multivariate logistic regression model was done starting with the variable with the strongest relationship with adherence to HIV testing guidelines among female sex workers. Other independent variables were added to this model one after another using forward stepwise method. The added variable would be dropped if it weakens the relationship between the first variable and the outcome. The stepwise logistic regression model was used to automatically select a reduced number of independent variables to build the best performing logistic regression model. The effect measure odds ratio was used to assess the relationship with adherence to the testing guidelines after adjusting for other covariates. Adjusting for other covariates was meant to take care of possible confounding by one independent variable on the relationship of another independent variable and study outcome, adherence. Factors with p-value ≤0.05 significance level were considered statically significant.

## RESULTS

### Socio-demographic characteristics of the study participants

Most participants were aged 25-30 years (43%; n=85, and nearly (51%; n=100) joined the sex work industry around the age of 18 to 24 years. The main source of income for this study sample was sex work (64%; n=128). Majority were living in their own or rented homes (89%; n=171), had children (86%; n=171) and most of them had been to primary school (49%; n=97) and secondary school (41%; n=82) respectively (Table 1).

### Adherence to the HIV testing guidelines and associated characteristics

As shown in table 2, (89.5%; n= 179) of the HIV negative participants reported that they tested for HIV infection in the last twelve months, (56%; n=100) of these had tested ≥3 times. The proportion of adherence to the HIV testing guidelines were significantly higher among the HIV-negative FSWs who attained secondary education than among the FSWs who had obtained primary or had no education at all (53% vs. 37%, p=0.03). Of the 18 FSWs in the study who received a complete FSWs minimum health package in the past 3 months, they all adhered to the testing guidelines. (100% vs 0% p=0.001).

### Factors that facilitated adherence to the HIV testing guidelines

There were several factors that participants highlighted as potential motivating factors. The desire to know their HIV status and seek treatment early should they be found to be HIV positive was prime (41%; n=111). Other (11%; n=32) cited easy accessibility to testing services since the testing services were brought nearer to their communities; while (13%; n=37) indicated that their perception of being at a high risk of HIV infection due to condom bursting during sexual intercourse prompted them to seek HIV testing. Furthermore, (10%; n=27) of the participants indicated that illnesses were another reason that led to take up regular HIV testing **(Figure 1)**.

**Figure 1:**
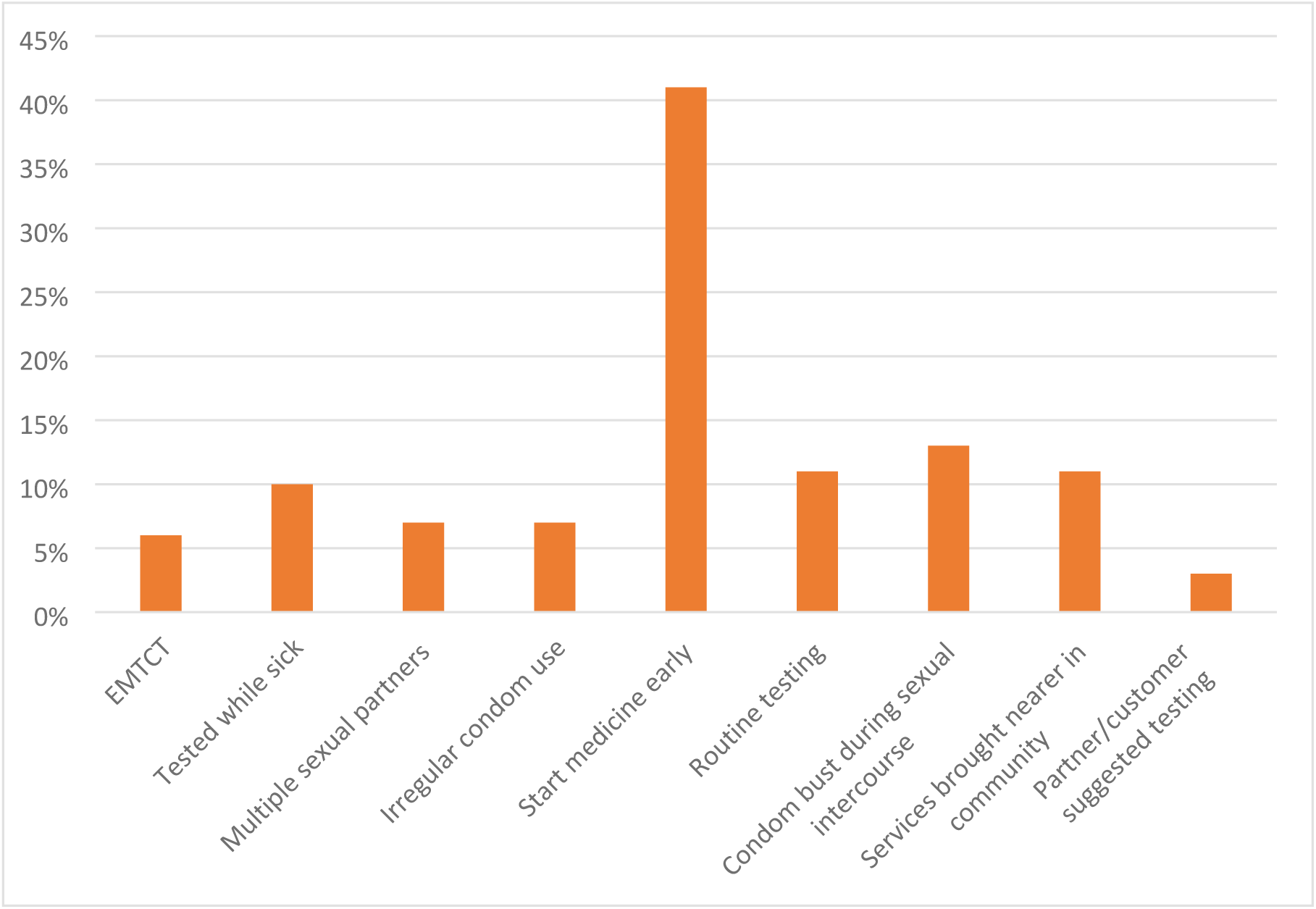
Factors that facilitated adherence to the HIV testing guidelines.

#### Factors associated with non-adherence to the HIV testing guidelines

Approximately (22%; n= 40) FSWs reported challenges to routine HIV testing including perception of risk of HIV infection and fear of knowing their HIV status (43%; n=17), lack of transport to visit a health facility for regular HIV testing (28%; n=11), and fear of being stigmatized by the health workers (13%; n=5). Although, (64% n=114) of the FSWs were aware about the testing guidelines, out of those who knew the guidelines, only (60%; n=60) were adherent, (48% n=26) of the participants indicated they were consistently using condoms and did not see the reason of routine testing, and that the testing centres opening hours are not work friendly to FSWs (22%; n=12) and others were just reluctant to test due to unknown reasons (18%; n=10) **(Figure 2**).

**Figure 2:**
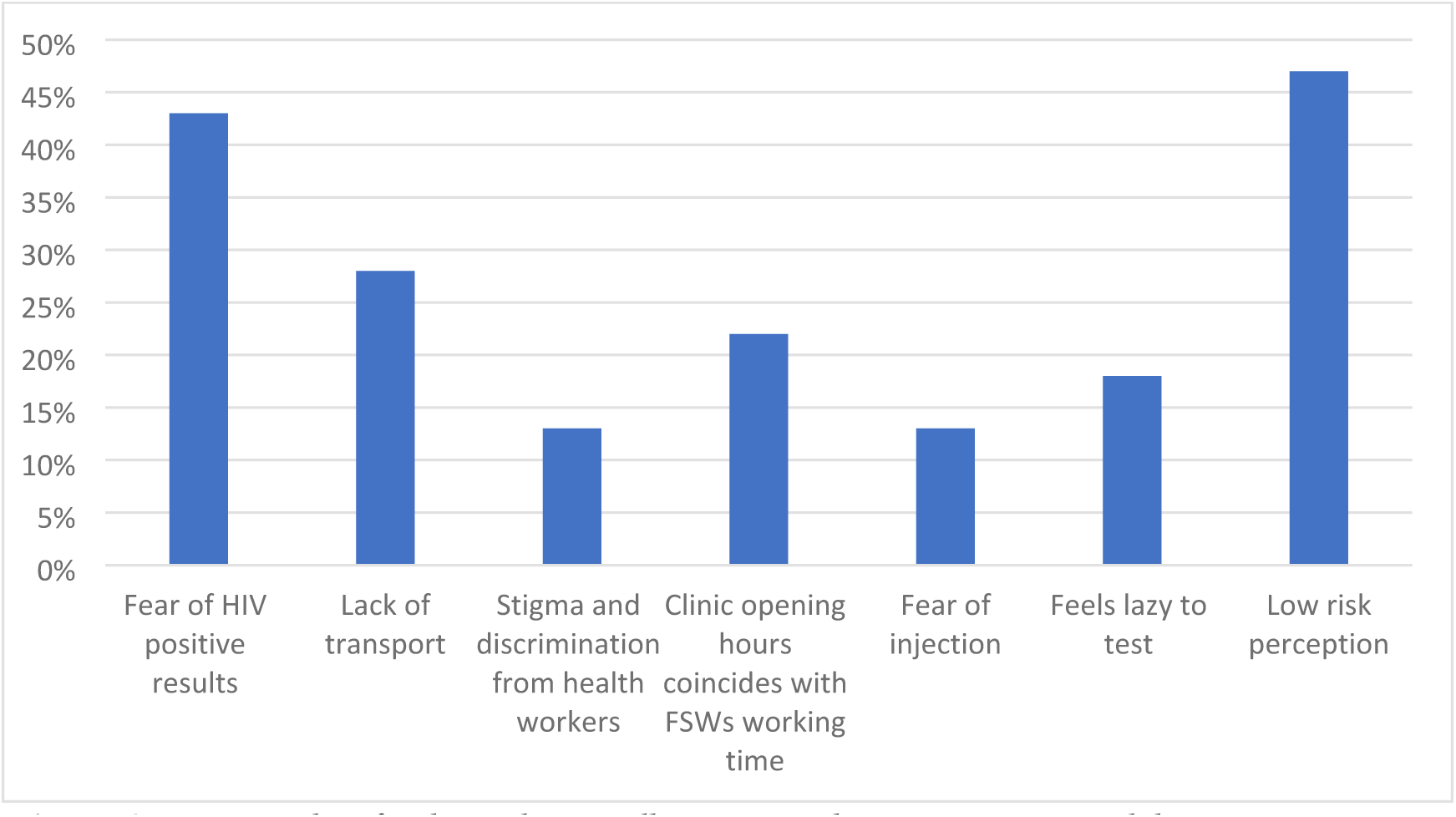
Factors that facilitated non-adherence to the HIV testing guidelines.

### Univariate Analysis of factors associated with adherence to the HIV testing guidelines among HIV-negative FSW

On univariate analysis (Table 3) participants who had attained secondary education were more likely to adhere to the testing guidelines than those that had attained primary or no education at all (OR=1.94, 95% CI: 1.06 - 3.55, p=0.031). Those that had tested for STI in the last three months (OR= 2.13, 95% CI: 0.95 - 4.74, p=0.065 as well as those who had accessed HIV testing in a drop-in centre (OR= 5.90, 95% CI: 0.71 - 49.1, p=0.101) had higher odds of adhering to the testing guidelines

**Table 3:** Univariate analysis of factors associated with adherence to the HIV testing guidelines

| Factors | Univariate Analysis<br>OR (95% CIs) | p-value |
| --- | --- | --- |
| **Age** |  |  |
| 8-24 | 1 |  |
| 25-30 | 0.67(0.33;1.35) | 0.258 |
| 31-49 | 1.02(0.46;2.24) | 0.966 |
| **Education** |  |  |
| Primary | 1 |  |
| ≥Secondary | 1.94 (1.06; 3.55) | 0.031 |
| **Marital status** |  |  |
| Single/Never married | 1 |  |
| Married and cohabiting | 0.58(0.19;1.77) | 0.336 |
| Separated/Divorced/Widowed | 0.68(0.33;1.41) | 0.304 |
| **Another Paying job** |  |  |
| No |  |  |
| Yes | 0.99(0.53;1.82) | 0.962 |
| **Condom use** |  |  |
| No |  |  |
| Yes | 1.17(0.47;2.92) | 0.732 |
| **Drug use** |  |  |
| No |  |  |
| Yes | 1.54(0.80;2.97) | 0.197 |
| **Used alcohol** |  |  |
| No |  |  |
| Yes | 1.26(0.61;2.60) | 0.535 |
| **STI screening last three months** |  |  |
| No |  |  |
| Yes | 2.13(0.95;4.74) | 0.065 |
| **Routine medical check-up** |  |  |
| No |  |  |
| Yes | 0.86 (0.74;1.02) | 0.076 |
| **Knowledge of testing guidelines** |  |  |
| No |  |  |
| Yes | 1.00 (0.88; 1.14) | 0.989 |
| **Housing status** |  |  |
| Own/rented house | 1 |  |
| Housed by relatives/friends | 0.40 (0.14; 1.13) | 0.084 |
| No shelter | 1.27 (0.11; 14.5) | 0.847 |
| **HIV testing awareness** |  |  |
| No |  |  |
| Yes | 1.61(0.66; 3.95) | 0.297 |
| **Risk perception to HIV risk** |  |  |
| No risk | 1 |  |
| low risk | 1.17(0.47;2.88) | 0.738 |
| ≥High risk | 0.91(0.36;2.28) | 0.838 |
| **Social support** |  |  |
| No |  |  |
| Yes | 0.89(0.50;1.61) | 0.699 |
| **Use of drop-in centres** |  |  |
| No |  |  |
| Yes | 5.90 (0.71; 49.1) | 0.101 |

### Multivariate analysis of factors associated with adherence to the HIV testing guidelines among FSW

In the multivariate logistic regression analysis (Table 4), attaining a secondary education, was independently associated with adherence to the HIV testing guidelines, after adjusting for use of drop-in centres (DIC) and routine medical check-up (OR: 1.86, 95% CI: 1.01 - 3.44, p=0.047).

**Table 4:** Multivariate analysis of factors associated with adherence to the HIV testing guidelines among FSW

| Factors | Multivariate Analysis<br>OR (95% CIs) | p-value |
| --- | --- | --- |
| **Age** |  |  |
| 8-24 | 1 |  |
| 25-30 | 0.58 (0.22;1.53) | 0.270 |
| 31-49 | 1.12(0.40;3.20) | 0.832 |
| **Education** |  |  |
| Primary | 1 |  |
| ≥Secondary | 2.18 (1.01; 4.71) | 0.047 |
| **Routine medical check-up** |  |  |
| No |  |  |
| Yes | 0.88 (0.73; 1.05) | 0.167 |
| **Housing status** |  |  |
| Own/rented house | 1 |  |
| Housed by relatives/friends | 0.37 (0.12; 1.13) | 0.081 |
| No shelter | 0.83 (0.06; 11.1) | 0.886 |

## Discussion

In this study we examined the extent to which HIV negative FSWs adhere to the testing guidelines by measuring the frequency of HIV testing in the year preceding the survey, and the associated factors. The HIV testing guidelines in Uganda, indicate that FSWs should test every three months (5). However, in this study the researcher’s definition considers FSWs to have adhered to the testing guidelines if they tested for HIV at least three times in the last one year regardless of the interval.

On the assessment of testing frequencies as measure of adherence to the testing guidelines, the FSWs that had tested three or more times in the year preceding the study constituted 57%. This outcome is too low to meet the WHO recommendation of HIV testing at intervals of every three to six months among FSWs (3) and Uganda testing guidelines for FSWs of testing every three months (5) and far below to meet the updated UNAIDS 95-95-95 targets for epidemic control by 2025 which aim for 95% of those living with HIV to know their status, 95% of those who know their status to be on treatment and 95% of those on treatment to be virally suppressed (34). Previously, few studies have assessed frequencies of testing, and instead a lot has been reported on the recent testing outcome (26). With the introduction of self-testing, studies have reported a potential benefit of increasing the frequency of HIV testing among FSWs (35, 36), this study therefore highlights a need for programs and studies to actively track complementary HIV testing outcomes including accessibility to HIV testing services, willingness to test and frequency of HIV testing among FSWs so that the data can be used for evidence based service delivery.

In this study we found that education was significantly associated with adherence to the HIV testing guidelines. This could be because individuals with secondary or higher education have increased capacity to interpret the guidelines, while those with primary level or no education at all could affect comprehension to understand but also to appreciate the benefits of testing for HIV on a routine basis. Other studies have reported similar findings although in different populations and study outcomes. For example, a study on HIV testing among adolescents in Uganda found that individuals who had secondary or higher education were more likely to have ever tested than those who had no education at all (37). Similarly, an analysis of demographic and healthy survey in Zambia found that higher education attainment was a strong predictor of uptake of HIV testing among women of child bearing age (38). This finding is not isolated for Sub-Saharan African countries, as similar findings have also been reported in America (39). This implies that less educated FSWs may not fully understand the HIV testing guidelines and realize the benefits of adhering by following the recommended testing frequencies of every three months. Therefore, health promotion models for HIV testing with messaging specifically packaged for FSWs with no formal or primary education may help increase adherence to the HIV testing guidelines for HIV negative FSWs. In addition, equipping health workers, FSWs peer leaders and community educators with skills to deliver messaging in a simplified way may go a long way to improve adherence to the HIV testing guidelines by FSWs. Further, Uganda has a behaviour change implementing partner supported by PEPFAR whose mandate is to develop communication messaging through mass media and print and to bridge the gap of service uptake and adoption of key practices (33), therefore there is need for real-time monitoring for continuous program improvement and to continually refine understanding of what behaviour change communication models work, under what contextual circumstances. Re-evaluating the impact of the available messaging on adherence to testing guidelines by FSWs, including the extent of wide dissemination of information and the level of engagement of peers in information sharing and education is key. This helps to ascertain if there is need for revision in the way the messages are packaged and disseminated and if they suit the purpose to achieve optimal benefits.

Findings from this study show that FSWs who were staying with friends and relatives, compared to those who were renting houses on their own, were likely to have increased number of HIV tests in a year. Although non-significant, this finding supports previous reports of other studies about relationships between social support and increased uptake of HIV testing. In a systematic review on uptake of HIV testing and counselling among FSWs, it was found that FSWs who had a higher uptake of HIV testing are those who were married, had peers to remind them to test or were encouraged by employers (26). Other studies (40, 41) have also shown that FSWs who have social support are likely to seek a range of health services such as condom use and other reproductive health services. The more the diverse sources of social support, the more the FSWs are likely to seek care including routine HIV testing. This underscores the need for strengthened programmatic and sustained peer to peer support among female sex workers, an intervention that has been proven to support increased access to HIV services among FSWs (14, 42) yet largely dependent on external funding with little funds allocation from governments (43). As such, predictable program financing for sex work broader HIV prevention programs including financing community support systems and behaviour change communications where feasible, is a strong pillar for sustained support for FSWs HIV testing programs.

Similarly, this study found that FSWs who were conducting routine health check-up at health facilities and those who had tested for STI in the last three months preceding the survey showed improved adherence to the testing guidelines. This confirms findings from previous studies, that health seeking behaviour influences decisions to regularly take HIV tests since the testing services are mostly provided in the hospitals (44, 45). WHO recommends that FSWs screen for STIs every three months (46). According to service delivery guidelines for key populations including FSWs in Uganda, FSWs who visit health facilities for various health needs and are identified to be practicing sex work should also be offered STI screening and provider initiated HIV testing (5). If this guidance was strictly followed by health workers, the results for this outcome would likely be reflected in the increased numbers of FSWs that are testing for HIV every three months as well as screening for STIs. However, due to stigma and discrimination by health workers, a barrier that was reported in this study and in other previous studies (47, 48), FSWs may not freely self-identify to be practicing sex work and this might imply that there are missed opportunities of routinely testing FSWs whenever they visit health facilities to seek for their other health care needs.

In this study, use of drop-in centres to test for HIV had improved adherence to the testing guidelines, but the relationship was not significant. Due to health disparities and barriers to access HIV services including increased testing among FSW, WHO recommends setting up of safe spaces for FSWs that allows meaningful engagement of FSWs to determine their health priorities, and where appropriate, non-judgmental services can be provided (3). Previous studies have reported the impact of drop-in centres to increased access to HIV services including HIV testing in Uganda (9), Mozambique (49) and Malawi (50). However, drop-in centres have not been taken to scale, are scarce and therefore inaccessible to FSWs who would have otherwise desired to access HIV testing through DICs, an issue that was noted in this study. According to the PEPFAR report (unpublished, 2021), there are 46 accredited DICs in all regions of Uganda, but Ministry of Health is making steps to increase the number of DICs and has since developed guidelines for establishing and operating DICs (51) to guide processes and the needed capacity required when setting up DICs. Setting up of various DICs in all districts of Uganda and supporting them to provide FSWs appropriate services will go a long way to improve availability of FSWs friendly testing services and thus improved adherence to the testing guidelines among FSWs.

FSWs in Uganda face various barriers that affect routine HIV testing which eventually affect adherence to the testing guidelines (9, 48). In this study barriers related to health system and individual challenges were reported including stigma and discrimination from health workers, costs associated with transportation, and the inflexibility of clinic opening hours that do not take into consideration FSWs work contexts. These findings are consistent with reports in other studies in Uganda and other countries in the region (7, 14, 52). Other barriers reported in this study were individual perceptions related to hesitancy to take routine tests because of fear to receive the news of HIV positive results and others perceived themselves at low risk because they were consistently using condoms and did not see the reason of routine testing. These barriers have for a long time been reported in many studies in Sub-Saharan Africa (26, 53-55) and they continue to be reported, an indication for consistent targeted health awareness education as well as to periodically evaluate health promotion campaigns and models of dissemination to ensure that concerns or the knowledge gaps of intended beneficiaries are addressed.

This study has several limitations. The questions in this study including the testing frequency history depended on self-report and this might have caused recall bias or biases related to social desirability, leading to internal validity of the findings. In addition, the eligibility to this study required that a participant is HIV negative, and we depended on self-disclosure of HIV positive status, we therefore cannot rule out possible false information of participants seropositivity status thus introducing measurement bias. However, this could be a minimal downside since we did not inform gate keepers about this eligibility criteria and had developed a pseudo short questionnaire that the research team would transition to whenever a participant disclosed their HIV positive status. Further, due to the cross-sectional nature of the study design, there is inability to draw causal relationships but rather to only determine associations. Lastly, in this study, we used convenience sampling, and this could have introduced selection bias. The findings from this study therefore need to be interpreted with caution.

## Conclusion

Based on the findings in this study, recommendations that may contribute to improved adherence to the testing guidelines by FSWs are those that increase the reach to testing services, intensity, and impact of behaviour change communication messaging targeting sex work especially those with lower education. Attaining secondary education or better was associated with adherence to the testing guidelines. This calls for enhancement of awareness campaigns regarding HIV testing guidelines as a measure to improve HIV testing among FSWs. Also, there is need for programs to scale up interventions that aim to improve access to and frequency of HIV testing, for example self-testing programs need to be expanded to reach all FSWs. In addition, continuous, and greater use of real time program evaluation, to refine understanding of what behaviour change communication models work, under what contextual circumstances is very key. As such, evaluating the available HIV testing campaigns and ascertain that they adequately promote adherence to the testing guidelines and if need be, package the messages in a way that is easy to comprehend by FSWs with less education is paramount. Lastly, equipping health workers and FSWs peer leaders with skills to deliver messaging in a manner understandable by less educated FSWs may go a long way to improve adherence to the HIV testing guidelines by FSWs.

## Data Availability

Due to conditions of ethical approvals of research among key populations, we are unable to provide access to the full dataset on a public repository. However, we are willing to de-identify transcripts and make them available upon reasonable request. Interested persons should contact the corresponding author

## Acknowledgments

We acknowledge Ms. Doreen Bakeiha and Ms. Florence Namimbi, the HIV Prevention advisors at Infectious Disease Institute who were the link to the network organisations and for availing us lists of all mapped hotspots in Kampala. We extend our appreciation to study participants, the network organisations and FSWs peers who supported us reach the participants for data collection. We thank the research assistants for ensuring quality data was collected.

## Author contributions

**Lydia Atuhaire**; Conceptualization, methodology development of interview tools, writing – original draft, writing – review & editing. **Constance S Shumba;** Review of interview tools, methodology, writing review and editing. **Innocent Maposa and Lovemore Mapahla**; Formal analysis, methodology, writing – review & editing; **Peter S Nyasulu;** Review of interview tools, methodology, writing-review and editing. **All authors;** Read and approved the final manuscript

## References

1. World Health Organisation. Consolidated guidelines on the use of antiretroviral drugs for treating and preventing HIV infection: recommendations for a public health approach – 2nd ed. Geneva 2016.

2. World Health Organization. Differentiated Service Delivery for HIV: A Decision Framework for HIV Testing Services. Geneva. 2018.

3. World Health Organization. Consolidated guidelines on HIV prevention, diagnosis, treatment and care for key populations: Geneva, Switzerland; 2016.

4. Macdonald V, Verster A, Baggaley R. A call for differentiated approaches to delivering HIV services to key populations. Journal of the International AIDS Society. 2017;20:21658.

5. Ministry of Health. Toolkit for Differentiated HIV and other Services Delivery Models for Key Populations in Uganda. 2019.

6. Ministry of Health. National HIV Testing Services Policy and Implementation Guidelines. Ministry of Health of Uganda; 2016.

7. Wanyenze RK, Musinguzi G, Kiguli J, Nuwaha F, Mujisha G, Musinguzi J, et al. “When they know that you are a sex worker, you will be the last person to be treated”: Perceptions and experiences of female sex workers in accessing HIV services in Uganda. 2017;17(1):11.

8. Narasimhan M, Yeh PT, Haberlen S, Warren CE, Kennedy CE. Integration of HIV testing services into family planning services: a systematic review. Reproductive health. 2019;16(1):1–12.

9. Pande G, Bulage L, Kabwama S, Nsubuga F, Kyambadde P, Mugerwa S, et al. Preference and uptake of different community-based HIV testing service delivery models among female sex workers along Malaba-Kampala highway, Uganda, 2017. BMC Health Services Research. 2019;19(1):1–11.

10. Mulongo S, Kapila G, Hatton T, Canagasabey D, Arney J, Kazadi T, et al. Applying innovative approaches for reaching men who have sex with men and female sex workers in the Democratic Republic of Congo. JAIDS Journal of Acquired Immune Deficiency Syndromes. 2015;68:S248–S51.

11. Ma H, Wang L, Gichangi P, Mochache V, Manguro G, Musyoki HK, et al. Venue-based HIV testing at sex work hotspots to reach adolescent girls and young women living with HIV: a cross-sectional study in Mombasa, Kenya. Journal of acquired immune deficiency syndromes (1999). 2020;84(5):470.

12. Bassett IV, Regan S, Mbonambi H, Blossom J, Bogan S, Bearnot B, et al. Finding HIV in hard to reach populations: mobile HIV testing and geospatial mapping in Umlazi township, Durban, South Africa. AIDS and Behavior. 2015;19(10):1888–95.

13. Cowan FM, Chabata ST, Musemburi S, Fearon E, Davey C, Ndori-Mharadze T, et al. Strengthening the scale-up and uptake of effective interventions for sex workers for population impact in Zimbabwe. Journal of the International AIDS Society. 2019;22:e25320.

14. Atuhaire L, Adetokunboh O, Shumba C, Nyasulu PS. Effect of community-based interventions targeting female sex workers along the HIV care cascade in sub-Saharan Africa: a systematic review and meta-analysis. Systematic Reviews. 2021;10(1):1–20.

15. Kelvin EA, George G, Mwai E, Kinyanjui S, Romo ML, Odhiambo JO, et al. A randomized controlled trial to increase HIV testing demand among female sex workers in Kenya through announcing the availability of HIV self-testing via text message. AIDS and Behavior. 2019;23(1):116–25.

16. Ndori-Mharadze T, Fearon E, Busza J, Dirawo J, Musemburi S, Davey C, et al. Changes in engagement in HIV prevention and care services among female sex workers during intensified community mobilization in 3 sites in Zimbabwe, 2011 to 2015. Journal of the International AIDS Society. 2018;21:e25138.

17. Wulandari LPL, Kaldor J, Januraga PP. High condom use but low HIV testing uptake reported by men who purchase sex in Bali, Indonesia. AIDS Care. 2018;30(10):1215–22.

18. Deering KN, Montaner J, Chettiar J, Jia J, Ogilvie G, Buchner C, et al. Successes and gaps in uptake of regular, voluntary HIV testing for hidden street-and off-street sex workers in Vancouver, Canada. AIDS Care. 2015;27(4):499–506.

19. Batona G, Gagnon M-P, Simonyan DA, Guedou FA, Alary M. Understanding the intention to undergo regular HIV testing among female sex workers in Benin: a key issue for entry into HIV care. JAIDS Journal of Acquired Immune Deficiency Syndromes. 2015;68:S206–S12.

20. Bengtson AM, L’Engle K, Mwarogo P, King’ola N. Levels of alcohol use and history of HIV testing among female sex workers in Mombasa, Kenya. AIDS Care. 2014;26(12):1619–24.

21. Setlhare K, Manyeagae GD. Factors associated with HIV testing among female sex workers in Botswana. Journal of AIDS and HIV Research. 2017;9(2):42–51.

22. Hladik W BA, Serwadda D, Tappero JW, Kwezi, R, Namakula DK, et al. Burden and characteristics of HIV infection among female sex workers in Kampala, Uganda–a respondent-driven sampling survey. BMC Public Health. 2017;17(1):565.

23. Muhindo R, Castelnuovo B, Mujugira A, Parkes-Ratanshi R, Sewankambo NK, Kiguli J, et al. Psychosocial correlates of regular syphilis and HIV screening practices among female sex workers in Uganda: a cross-sectional survey. AIDS Research and Therapy. 2019;16(1):1–9.

24. Martins TA, Kerr L, Macena RHM, Mota RS, Dourado I, Brito AMd, et al. Incentivos e barreiras ao teste de HIV entre mulheres profissionais do sexo no Ceará. Revista de Saúde Publica. 2018;52:64.

25. Damacena GN, Szwarcwald CL, de Souza Júnior PR, Dourado I. Risk factors associated with HIV prevalence among female sex workers in 10 Brazilian cities. JAIDS Journal of Acquired Immune Deficiency Syndromes. 2011;57:S144–S52.

26. Tokar A, Broerse JE, Blanchard J, Roura M. HIV testing and counseling among female sex workers: a systematic literature review. AIDS and Behavior. 2018;22(8):2435–57.

27. Decker MR, Wirtz AL, Pretorius C, Sherman SG, Sweat MD, Baral SD, et al. Estimating the impact of reducing violence against female sex workers on HIV epidemics in Kenya and Ukraine: a policy modeling exercise. American Journal of Reproductive Immunology. 2013;69:122–32.

28. Nyblade L, Reddy A, Mbote D, Kraemer J, Stockton M, Kemunto C, et al. The relationship between health worker stigma and uptake of HIV counseling and testing and utilization of non-HIV health services: the experience of male and female sex workers in Kenya. AIDS Care. 2017;29(11):1364–72.

29. Lillie TA, Persaud NE, DiCarlo MC, Gashobotse D, Kamali DR, Cheron M, et al. Reaching the unreached: performance of an enhanced peer outreach approach to identify new HIV cases among female sex workers and men who have sex with men in HIV programs in West and Central Africa. PloS One. 2019;14(4):e0213743.

30. Olawore O, Astatke H, Lillie T, Persaud N, Lyons C, Kamali D, et al. Peer Recruitment Strategies for Female Sex Workers Not Engaged in HIV Prevention and Treatment Services in Côte d’Ivoire: Program Data Analysis. JMIR public health and surveillance. 2020;6(4):e18000.

31. Shokoohi M, Noori A, Karamouzian M, Sharifi H, Khajehkazemi R, Fahimfar N, et al. Remaining gap in HIV testing uptake among female sex workers in Iran. AIDS and Behavior. 2017;21(8):2401–11.

32. Van Bulck L, Wampers M, Moons P. Research Electronic Data Capture (REDCap): tackling data collection, management, storage, and privacy challenges. European Journal of Cardiovascular Nursing. 2021.

33. FHI 360. Social and behavior change communication in Uganda. Providing strategic support to improve health program outcomes 2020.

34. UNAIDS. 2030-Ending the AIDS Epidemic, the 2025 AIDS Targets. https://www.unaidsorg/sites/default/files/2025-AIDS-Targets_enpdf Accessed December 27, 2021. 2021.

35. Wang C, Wang Y-J, Tucker JD, Xiong M-Z, Fu H-Y, Smith MK, et al. Correlates of HIV self-testing among female sex workers in China: implications for expanding HIV screening. Infectious diseases of poverty. 2020;9(1):1–9.

36. Shava E, Manyake K, Mdluli C, Maribe K, Monnapula N, Nkomo B, et al. Acceptability of oral HIV self-testing among female sex workers in Gaborone, Botswana. PloS One. 2020;15(7):e0236052.

37. Purba FD, Hunfeld JA, Fitriana TS, Iskandarsyah A, Sadarjoen SS, Busschbach JJ, et al. Living in uncertainty due to floods and pollution: the health status and quality of life of people living on an unhealthy riverbank. BMC Public Health. 2018;18(1):1–11.

38. Muyunda B, Musonda P, Mee P, Todd J, Michelo C. Educational attainment as a predictor of HIV testing uptake among women of child-bearing age: analysis of 2014 demographic and health survey in Zambia. Frontiers in public health. 2018;6:192.

39. Onyeabor OS, Iriemenam N, Adekeye OA, Rachel SA. The effect of educational attainment on HIV testing among African Americans. Journal of Health Care for the Poor and Underserved. 2013;24(3):1247–56.

40. Qiao S, Li X, Zhang C, Zhou Y, Shen Z, Tang Z. Social support and condom use among female sex workers in China. Health Care for Women International. 2015;36(7):834–50.

41. Mizinduko M, Moen K, Pinkowski Tersbøl B, Likindikoki SL, Alexander Ishungisa M, Leyna GH, et al. HIV testing and associated factors among female sex workers in Tanzania: approaching the first 90% target? AIDS Care. 2021:1–9.

42. Nnko S KE, Nyato D, Drake M, Casalini C, Shao A, Komba A, Baral S, Wambura M, Changalucha J. Determinants of access to HIV testing and counselling services among female sex workers in sub-Saharan Africa: a systematic review. BMC Public Health. 2019;19(1):15.

43. Kerrigan D, Kennedy CE, Morgan-Thomas R, Reza-Paul S, Mwangi P, Win KT, et al. A community empowerment approach to the HIV response among sex workers: effectiveness, challenges, and considerations for implementation and scale-up. The Lancet. 2015;385(9963):172–85.

44. Musekiwa A, Bamogo A, Shisana O, Robsky K, Zuma K, Zungu NP, et al. Prevalence of self-reported HIV testing and associated factors among adolescent girls and young women in South Africa: results from a 2017 nationally representative population-based HIV survey. Public Health in Practice. 2021;2:100093.

45. Nangendo J, Katahoire AR, Armstrong-Hough M, Kabami J, Obeng-Amoako GO, Muwema M, et al. Prevalence, associated factors and perspectives of HIV testing among men in Uganda. PloS One. 2020;15(8):e0237402.

46. World Health Organization. Global health sector strategy on sexually transmitted infections 2016-2021: towards ending STIs. World Health Organization; 2016.

47. Nakanwagi S, Matovu JK, Kintu BN, Kaharuza F, Wanyenze RK. Facilitators and barriers to linkage to hiv care among female sex workers receiving hiv testing services at a community-based organization in Periurban Uganda: A Qualitative Study. Journal of sexually transmitted diseases. 2016;2016.

48. Wanyenze RK, Musinguzi G, Kiguli J, Nuwaha F, Mujisha G, Musinguzi J, et al. “When they know that you are a sex worker, you will be the last person to be treated”: perceptions and experiences of female sex workers in accessing HIV services in Uganda. BMC International Health and Human Rights. 2017;17(1):1–11.

49. Lafort Y, Geelhoed D, Cumba L, Lázaro CdDM, Delva W, Luchters S, et al. Reproductive health services for populations at high risk of HIV: Performance of a night clinic in Tete province, Mozambique. BMC Health Services Research. 2010;10(1):1–9.

50. Vu L, Zieman B, Muula A, Samuel V, Tenthani L, Chilongozi D, et al. Assessment of community-based ART service model linking female sex workers to HIV care and treatment in Blantyre and Mangochi, Malawi. 2021.

51. Ministry of Health. Guidelines for establishing and operating drop-in centres for key populations in Uganda 2020.

52. Lafort Y, Greener L, Lessitala F, Chabeda S, Greener R, Beksinska M, et al. Effect of a ‘diagonal’intervention on uptake of HIV and reproductive health services by female sex workers in three sub-Saharan African cities. Tropical Medicine and International Health. 2018;23(7):774–84.

53. Aho J, Nguyen VK, Diakité S, Sow A, Koushik A, Rashed S. High acceptability of HIV voluntary counselling and testing among female sex workers: impact of individual and social factors. HIV Medicine. 2012;13(3):156–65.

54. Ameyan W, Jeffery C, Negash K, Biruk E, Taegtmeyer M. Attracting female sex workers to HIV testing and counselling in Ethiopia: a qualitative study with sex workers in Addis Ababa. African Journal of AIDS Research. 2015;14(2):137–44.

55. Chanda MM, Perez-Brumer AG, Ortblad KF, Mwale M, Chongo S, Kamungoma N, et al. Barriers and facilitators to HIV testing among Zambian female sex workers in three transit hubs. AIDS Patient Care and STDS. 2017;31(7):290–6.

